# Study protocol for a randomised controlled trial investigating the effects of Mindfulness Based Stress Reduction on stress regulation and associated neurocognitive mechanisms in stressed university students: The MindRest study

**DOI:** 10.1101/2023.05.24.23290218

**Authors:** Nikos Kogias, Dirk E. M. Geurts, Florian Krause, Anne E. M. Speckens, Erno J. Hermans

## Abstract

**Background:** Stress-related disorders are a growing public health concern. While stress is a natural and adaptive process, chronic exposure to stressors can lead to dysregulation and take a cumulative toll on physical and mental well-being. One approach to coping with stress and building resilience is through Mindfulness-Based Stress Reduction (MBSR). By understanding the neural mechanisms of MBSR, we can gain insight into how it reduces stress and what drives individual differences in treatment outcomes. This study aims to establish the clinical effects of MBSR on stress regulation in a population that is susceptible to develop stress-related disorders (i.e., university students with mild to high self-reported stress), to assess the role of large-scale brain networks in stress regulation changes induced by MBSR, and to identify who may benefit most from MBSR.

**Methods:** This study is a longitudinal two-arm randomised, wait-list controlled trial to investigate the effects of MBSR on a preselected, Dutch university student population with elevated stress levels. Clinical symptoms are measured at baseline, post-treatment, and three months after training. Our primary clinical symptom is perceived stress, with additional measures of depressive and anxiety symptoms, alcohol use, stress resilience, positive mental health, and stress reactivity in daily life. We investigate the effects of MBSR on stress regulation in terms of behaviour, self-report measures, physiology, and brain activity. Repetitive negative thinking, cognitive reactivity, emotional allowance, mindfulness skills, and self-compassion will be tested as potential mediating factors for the clinical effects of MBSR. Childhood trauma, personality traits and baseline brain activity patterns will be tested as potential moderators of the clinical outcomes.

**Discussion:** This study aims to provide valuable insights into the effectiveness of MBSR in reducing stress-related symptoms in a susceptible student population and crucially, to investigate its effects on stress regulation, and to identify who may benefit most from the intervention.

**Trial registration:** Registered on September 15, 2022, at clinicaltrials.gov, NCT05541263. https://clinicaltrials.gov/ct2/show/study/NCT05541263

## Background

Stress-related mental disorders, such as anxiety, depression, and post-traumatic stress disorder are a significant public health concern with substantial and often debilitating mental and physical symptoms (1). In 2017, 540 million people were affected world-wide by anxiety and depression alone (GBD). These disorders contribute to shorter life expectancy (3), lower quality of life (4), as well as a large economic burden to society (5). As the term “stress-related disorders” suggests, a common component in the development of such disorders is (chronic) exposure to stressors. These can be internal or external factors creating a (perceived) threat to one’s well-being. Exposure to stressors can prompt a response (i.e. the stress response) that includes a complex set of psychological, cognitive, neural, metabolic, immunological, and physiological changes developed to aid individuals to better adapt to challenges in their life (6). However, chronic exposure to stressors can lead to a dysregulation of these complex interactions causing a cumulative toll on physical and mental well-being, described as allostatic overload (6,7).

Perceived stress is a robust predictor of the development of psychopathology, and stress-regulation processes have been shown to play a protective role in that respect (8). Therefore, adequate coping with stress may substantially contribute to stressnresilience, preventing the deterioration of mental health. An effective approach with this very aim is Mindfulness Based Stress Reduction (MBSR) (9).

Introduced into clinical practice in 1979 by Kabat-Zinn, mindfulness is defined as non-judgemental present-moment, intentional awareness. Mindfulness training thus promotes deliberately paying attention with a curious, open attitude to one’s habitual affective, cognitive, and behavioural reactions to stress, ultimately learning to deal more effectively with stressful situations (10,11). This intervention has been shown to be effective in reducing symptoms of perceived stress, anxiety, and depression in healthy and clinical populations (9,12). However, these interventions are not universally effective, and there is a growing recognition for the need to understand individual differences in treatment response.

A mechanistic understanding of stress regulation via mindfulness could shed light on *how* MBSR reduces stress and *what* drives individual differences in treatment outcome. Research into psychological mechanisms of MBSR and derivatives of this program, described as Mindfulness-Based Interventions (MBIs), has shown that self-reported mindfulness skills and decentring play a mediating role in stress and anxiety symptom reduction (13,14). Other potential mechanisms include different emotion regulation processes, reducing ruminative thoughts, and acceptance, suggesting that mindfulness can influence affective processing (15). These mechanisms provide a framework for stress regulation that might be considered different to other common stress management strategies, such as cognitive reappraisal, which depend mainly on enhancing executive control (16). Additionally, there is some evidence that MBIs can influence different cognitive processes (15,17). Various attention-related and working memory-related functions have been shown to be affected following a MBI, although evidence is limited (18,19).

In line with psychological research, research on the neural mechanisms of MBIs provides preliminary evidence on the involvement of neural structures engaged in both cognitive and affective processing (20). Some of the most robust findings have included structural and functional changes in brain regions such as the insula, the amygdala, the anterior cingulate cortex (ACC), and multiple prefrontal cortex (PFC) regions (e.g., dorsolateral PFC and ventromedial PFC) (20,21). Interestingly, these regions are considered core nodes of three large-scale brain networks, which are thought to play a role in the stress response as well as in the development of various stress-related disorders (22–24). First, the salience network (SN) is considered to be central in affective processes integrating relevant autonomic, interoceptive and emotional information (25). Second, the executive control network (ECN) is involved in higher-order cognitive functions, such as goal-directed problem solving, top-down control, and decision making (26). Third, the default mode network (DMN) is associated with self-referential and social-cognitive processing (27). In addition, a recent review of accumulating data suggests that mindfulness is specifically related to functional connectivity changes in these networks (28). Therefore, the clinical effects of MBSR could potentially be driven by a change in these networks’ configurations under stress, constituting a more adaptive stress-regulation.

In the current study we focus on how MBSR can affect different aspects of stress regulation in terms of behaviour, self-report measures, physiology and brain activity, across two orthogonal dimensions. The first dimension covers stress-regulation processes spanning from regulation directly related to external stressors (exogenously driven regulation) to regulation related to internal stressors, such as ruminative thoughts, or stressed states not directly related to external stressors, e.g., in the aftermath of acute stress (endogenously driven regulation). The second dimension spans from stress-related processes requiring active, goal-directed responses (i.e., explicit regulation) to stress-related processes when no action is required (i.e., implicit regulation). This framework yields four domains of stress-regulation processes. For example, exogenous-explicit regulation occurs while trying to deliberately apply emotional regulation during a stressful event such as an exam. Exogenous-implicit regulation describes processes like automatic reactions to fearful stimuli such as the anticipation of a sharp pain. Endogenous-explicit regulation involves processes like deliberate reappraisal of intrusive thoughts. An example of endogenous-implicit regulation is one’s automatic reactions to anxious internal states (e.g., rumination).

We apply this framework in the design of the current longitudinal randomised controlled trial, whereby we investigate the effects of MBSR on a preselected, Dutch, university-student population with high self-reported levels of perceived stress.

University students have been shown to be a population at risk of developing stress-related symptoms. Frequent examination periods, assignment deadlines, restricted financial support, time pressure to complete studies, and future uncertainty are some of the main sources of prolonged stress for students (29). This may have implications for academic performance, private life, and overall health, thus leading to increased personal and societal burden. There is already evidence for a beneficial effect of MBSR on stress in general student samples (9). However, studies targeting specifically highly stressed students, those presumably most at risk for mental health problems, are lacking. This is a population that may greatly benefit from enhanced stress-regulation potentially preventing stress-related symptomatology. It is therefore key to establish whether MBSR is indeed effective in reducing stress in this specific student population.

Thus, the goals of the current study are to (1) establish clinical effects of MBSR in a population of Dutch university students with high levels of perceived stress, (2) identify the effect of MBSR on stress regulation, (3) assess the role of large-scale networks in this process, and (4) identify for whom MBSR might be more beneficial. Our main hypothesis regarding aim (1) is that MBSR is effective at reducing self-reported perceived stress in a population of highly stressed university students, sustained over a 3-month follow-up period. We expect to replicate previous studies showing the efficacy of MBSR within student populations (30–33) and we expect that MBSR is also effective at reducing other relevant clinical symptoms in this population, such as depression and anxiety symptoms (9). Regarding aims (2) and (3) (i.e., the effects of MBSR on stress regulation and the role of large-scale networks in this process), we hypothesise that MBSR leads specifically to concurrent enhancement of cognitive control *and* affective processing across the different domains of stress-regulation described above. Moreover, we expect that these measures at baseline will be important in predicting individual differences in effectiveness of MBSR, and that changes in these measures will be accompanied by changes at the clinical level for aim (4).

## Methods

### Design

This study is a two-arm randomised, wait-list controlled trial with ethical approval from the local medical-ethical committee (METC Oost-Nederland). Participants are randomised into a treatment and a wait-list control group after baseline measurements including clinical, neurocognitive, and ecological momentary assessments. In the following two months, the treatment group participates in an MBSR training, and the wait-list group receives no planned intervention for two months, serving the purpose of assessing the effect of the intervention on the study outcomes against not receiving treatment during the same time-period. Post-treatment measurements take place three months after baseline, and they include another set of clinical, neurocognitive, and ecological momentary assessments. Six months after baseline there is a follow-up clinical assessment. After this assessment, the participants in the wait-list group receive the MBSR training, after which they complete a final clinical assessment.

**Figure 1.**
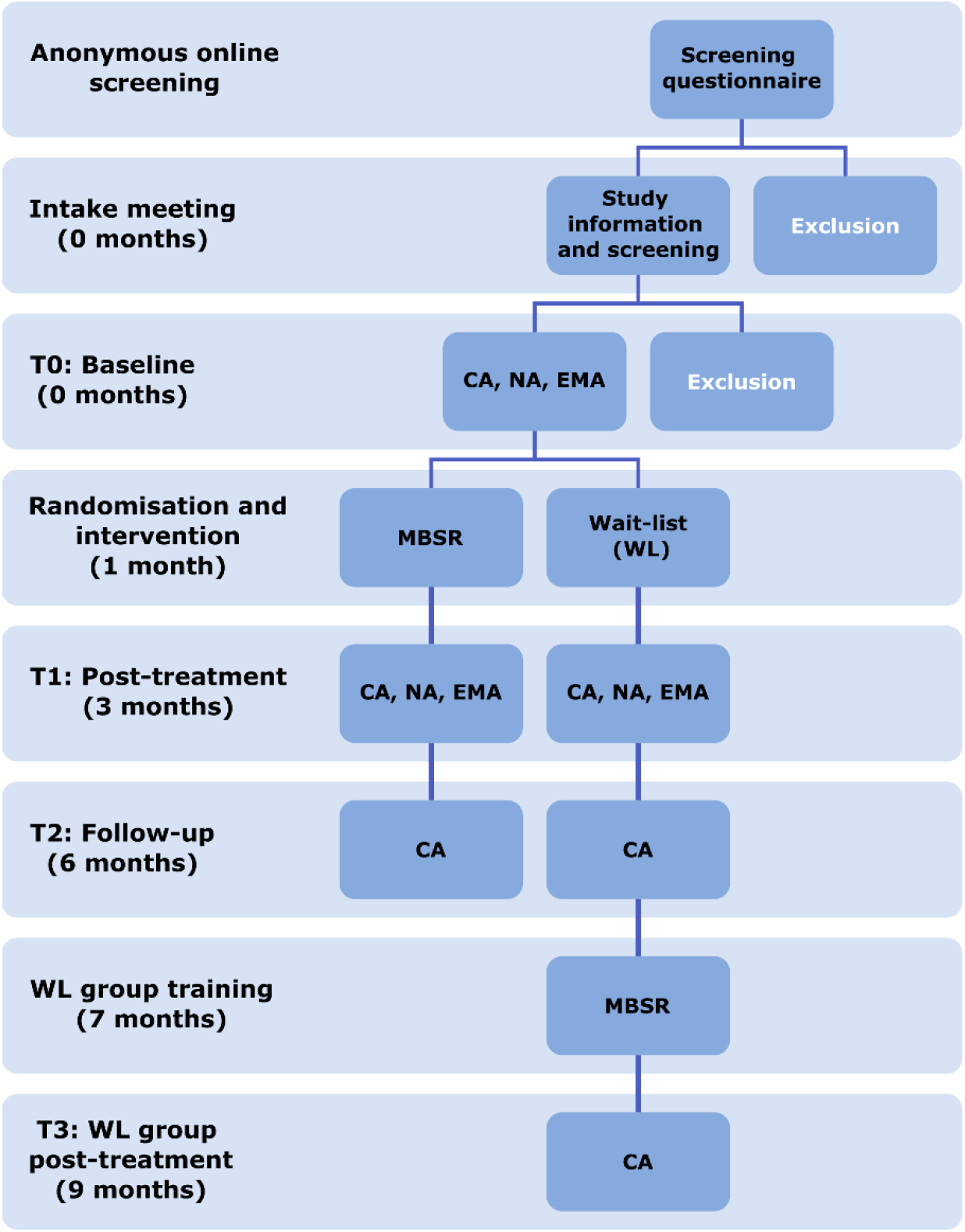
Study design. CA: clinical assessments, NA: Neurocognitive assessments, EMA: Ecological Momentary Assessments

### Population

We aim at recruiting 60 students per group (total: 120) from Radboud University, Radboudumc, and university of applied sciences Hogeschool Arnhem Nijmegen, with high perceived stress (see sample size calculation in analysis section). This target includes an expected attrition rate of 10%, leading to a total sample of 108 participants. We include students who are above 18 years, able to give consent, and who are mildly to highly stressed achieving a score ≥ 16 on the perceived stress scale (PSS) (34,35). Participants are excluded (1) if they are receiving current specialised psychological or psychiatric treatment or medication, (2) if they have insufficient comprehension of the Dutch language, (3) if they have physical, cognitive, or intellectual impairments interfering with participation, such as deafness, blindness, or sensorimotor handicaps, (4) if they were formerly or currently involved in a Mindfulness Based Stress Reduction or a Mindfulness Based Cognitive Therapy training, (5) if they have a current drug or alcohol addiction, and (6) if they have contraindications for MRI scanning (e.g., pacemaker, implanted metal parts, deep brain stimulation, claustrophobia, epilepsy, brain surgery, pregnancy).

### Intervention

The mindfulness intervention used in this study is an MBSR training based on the MBSR programme developed by Kabat-Zinn (1982) and has been widely investigated in the last few decades (9,36). The training consists of 8 weekly group sessions lasting 2,5 hours. A silent day of 6 hours is also included, as well as daily home practice assignments of a recommended 45 minutes. During the training participants learn to intentionally focus their attention on the present moment in an accepting and non-judgemental way, rather than ruminating about past and future experiences. The training includes formal exercises during which participants will practice the body scan, sitting meditation, walking meditation and mindful movement. Informal exercises are also included, such as performing a daily activity with full attention to the present experience. The training is led by qualified teachers meeting the advanced criteria of the Association of Mindfulness Based Teachers in the Netherlands and Flanders (www.vmbn.nl).

### Randomisation

Groups of maximally 20 participants at a time (because of limitations in scanning capacity) are recruited and are randomly assigned to either the MBSR or waitlist control condition. This ensures that equal numbers of experimental and control group participants are recruited at any given time-period. Randomisation is conducted after baseline clinical, neurocognitive, and ecological momentary assessments. The procedure is carried out by an automated custom Python script using computer-generated random numbers, taking into account stratification for gender and University (i.e., Radboud University, or Hogeschool Arnhem Nijmegen). Baseline measurements occur before group assignments and therefore both participants and researchers are blinded to these measurements. Post-treatment measurements are not blinded.

### Questionnaires (Clinical Assessments)

We measure perceived stress with the Perceived Stress Scale (PSS), a 10-item questionnaire (range 0-40), which evaluates the degree to which an individual perceives their life as unpredictable, uncontrollable, and overloading. This scale was found to have good internal consistency (*α=*0.84-0.86) and test-retest reliability (*rtt*=0.85) (34,35) Depressive symptoms are assessed with the Inventory of Depressive Symptomatology Self-Report (IDS-SR), which is a 30-item self-report measure of depressive symptom severity (range 0-84). This scale was found to have excellent internal consistency (*α=*0.93) (37,38).

Anxiety symptoms are assessed with the State and Trait Anxiety Inventory (STAI), which is a self-reported 20-item measure of trait (range 20-80) and a self-reported 20-item measure (range 20-80) state anxiety. The questionnaire was found to have good internal consistency for state (*α=*0.83-0.92) and trait (*α=*0.86-0.92). Test-retest reliability was good for trait (*rtt*=0.73-0.86) and poor for state (*rtt*=0.16-0.62) (39,40).

Alcohol use is assessed with the Alcohol Use Disorders Identification Test (AUDIT), which is a 10-item screening tool developed by the World Health Organization (WHO) to assess alcohol consumption, drinking behaviours, and alcohol-related problems (range 0-40). The questionnaire was found to have high internal consistency (*α=*0.80-0.86) and high test-retest reliability (*rtt*=0.84) (41–43).

Childhood trauma is assessed with the Maltreatment and Abuse Chronology of Exposure Scale (MACE-X), which is a self-reported questionnaire which is used to assess the extent as well as the severity of traumatic experiences of participants in their childhood. We use a Dutch translation of the original English questionnaire. The original questionnaire in English was found to have high test-retest reliability (*rtt*=0.91) (44).

Personality traits are assessed with the NEO Five Factor Inventory (NEO-FFI), which is a 60-item self-reported questionnaire and covers a set of five broad personality trait dimensions or domains: Extraversion, Agreeableness, Conscientiousness, Neuroticism, and Openness to Experience. The questionnaire was found to have high internal consistency for all subscales (*α=*0.74-0.89) and test-retest reliability (*rtt*=0.86-0.90) (45)

Repetitive negative thinking is assessed with the Perseverative Thinking Questionnaire (PTQ), which is a 15-item self-reported questionnaire and is used to assess repetitive negative thinking in a content-free manner (range 0-60). The questionnaire was found to have excellent internal consistency (*α=*0.94-0.95) and satisfactory test-retest reliability (*rtt*=0.69) (46).

Cognitive reactivity is assessed with the Leiden Index of Depression Sensitivity – Revised (LEIDS-R), which is a 34-item self-report questionnaire measuring cognitive reactivity to sadness (range 0-136). The questionnaire was found to have good internal consistency on all subscales (*α=*0.77-0.89) (47).

The concept of allowing of emotions is assessed with the Acceptance and Action Questionnaire (AAQ), which is a 10-item self-reported questionnaire measuring psychological flexibility and experiential acceptance (range 10-70). The questionnaire was found to have good internal consistency (*α=*0.84) and test-retest reliability (*rtt*=0.79-0.81) (48).

Mindfulness skills are assessed with the short version of the Five-Facet Mindfulness Questionnaire (FFMQ-SF), which is a 24-item self-reported questionnaire measuring five aspects of mindfulness, namely observation, description, aware actions, non-judgemental inner experience, and non-reactivity. The questionnaire was found to have good internal consistency for all subscales (*α=*0.73-0.91) (49).

Self-compassion is assessed with the short version of the Self-Compassion Scale (SCS-S), which is a 12-item self-report questionnaire consisting of six components, including self-kindness, self-judgment, common humanity, isolation, mindfulness, and over-identification. The questionnaire was found to have good internal consistency (*α=*0.86) (50).

Self-reported stress resilience is assessed with the Connor-Davidson Resilience Scale (CD-RISC), which is a 25-item questionnaire (range 0-100). The questionnaire was found to have good internal consistency (*α=*0.89) and test-retest reliability (*rtt*=0.87) (51).

Positive mental health is assessed with the short form of the Mental Health Continuum (MHC-SF), which is a 14-item self-report questionnaire that assesses emotional, psychological and social well-being (range 0-70). The questionnaire was found to have good internal consistency (*α=*0.89) and moderate test-retest reliability (*rtt*=0.65-0.70) (52).

All self-report questionnaire data are acquired using CastorEDC, an Electronic Data Capture program (https://www.castoredc.com/) during measurement visits.

### Stress-regulation tasks (Neurocognitive Assessments)

We assess these four domains of stress regulation (exogenous-explicit, exogenous- implicit, endogenous-explicit, and endogenous-implicit) using different outcome measures: self-report, behaviour, physiology, and brain imaging. Self-report questionnaires are used to assess perceived stress (e.g., subjective fear ratings).

These are acquired via Expyriment (53), the software we use to present task stimuli, along with assessments of behavioural responses (e.g., reaction time, response accuracy) recorded during stress regulation tasks. Physiological response assessments of autonomic nervous system measures (e.g., skin conductance, heart rate, and pupil size) are used to measure affective responses to stressors using BrainVision Recorder (Brain Products; Gliching, Germany) and Eyelink-1000 Plus eye-tracker (SR Research, Ottawa, Canada). We additionally use brain imaging to assess brain activity and connectivity within and between large-scale brain networks using a Siemens Skyra 3T MR system (Siemens, Erlangen, Germany). In addition, we record respiration via BrainVision Recorder (Brain Products; Gliching, Germany) during the neurocognitive tasks to correct the MR images for physiological noise. Saliva samples are acquired while performing the tasks to measure salivary cortisol levels as a response to stressors using salivette cotton swab tubes (SARSTEDT, Numbrecht, Germany).

**Figure 2.**
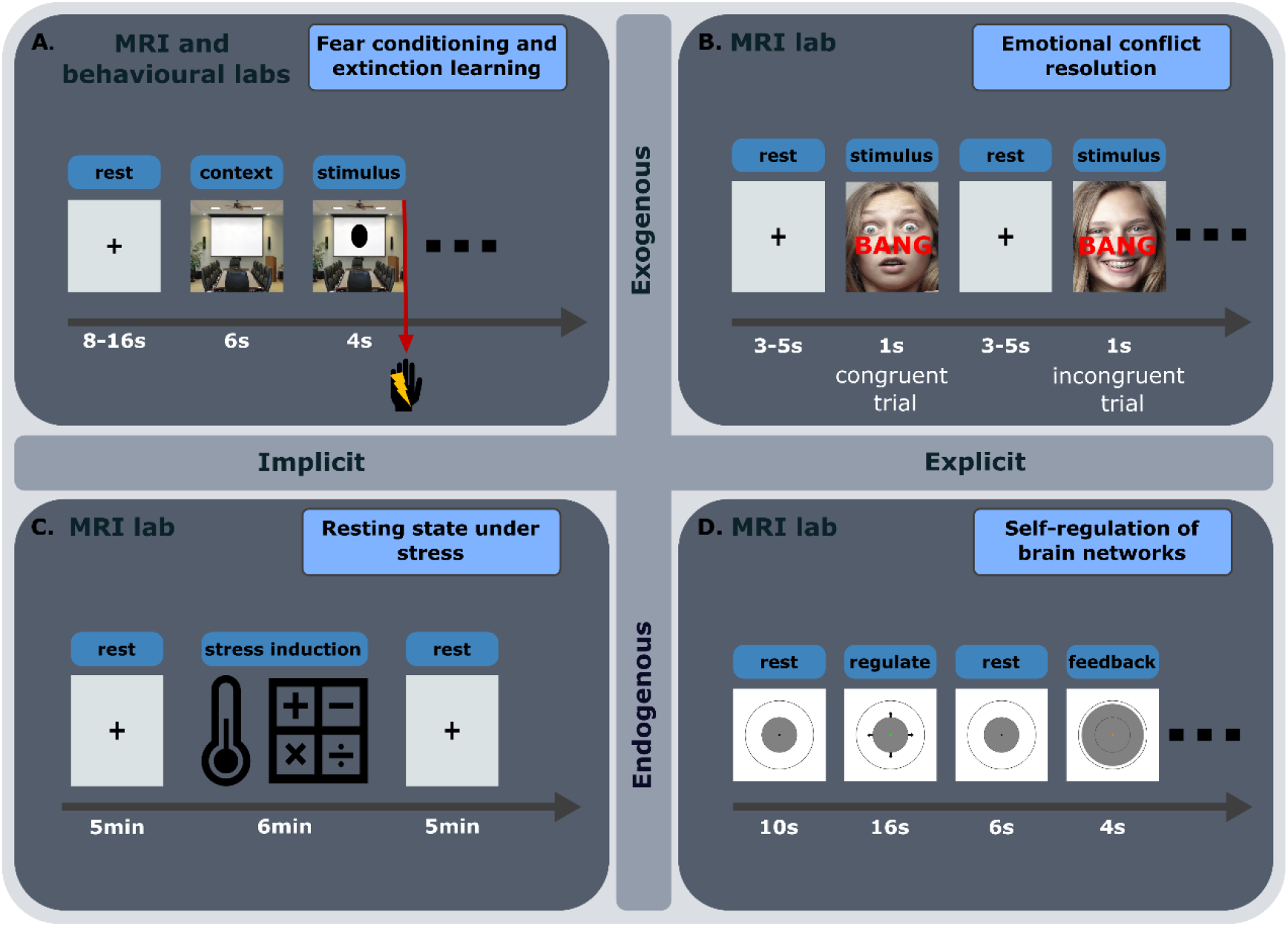
Experimental design for each of the tasks residing in four domains of stress regulation (implicit-exogenous, explicit-exogenous, implicit-endogenous, and explicit- endogenous) with example stimuli. A) An image of a room is shown on the screen as the context. Shortly after, a shape (e.g. circle or square) is shown within the room providing information on how likely it is to receive an electrical stimulation on the hand. During the conditioning phase, electrical stimulations are administered at the end of 35% of trials depicting one of the shapes. The other shape is never paired with an electrical stimulation. In the remaining phases of the task the same stimuli are shown but electrical stimulations are never administered. B) Participants select the emotion depicted on a face (happy or scared) while ignoring the word describing an emotion (happy or scared; “blij” or “bang” in Dutch) superimposed on top of the face. The word can either match the emotion on the face (congruent trial) or not (incongruent trial). Depicted stimuli(61) are examples; actual stimuli will be taken from Ekman and Friesen. C) Participants fixate their gaze on a fixation cross for 5 minutes during a resting state scan. A stress induction procedure follows, including a socially evaluated cold pressor test and a mental arithmetic task. Another 5-minute resting state scan is acquired after the stress induction procedure. D) Participants are asked to make the grey disk larger or smaller using only mental strategies. After a short rest they see the size of the disk change, reflecting their brain activity during their regulation period. This serves as feedback which can be used to learn and improve one’s strategies to control their brain activity.

#### Implicit, exogenous stress-regulation – Fear Conditioning and Extinction task

Implicit, exogenous stress regulation is assessed using a differential fear conditioning and extinction paradigm in the MRI scanner, since this task involves measuring spontaneous responses to an external stressor. Mild electrical stimulation to the fingers is used as the unconditioned stimulus (US). Two shapes are used as conditioned stimuli (CS), and they are presented for 4s on top of a specific background image (the same for both stimuli) with an average intertrial interval of 12s. One of the two shapes is sometimes paired with a US during the final 200ms of the stimulus presentation (CS+) and the other one is never paired with the US (CS-). Reinforcement rate using the US is set at 35%. After fear conditioning has occurred, we proceed with a fear extinction task inside the MRI scanner. The two CS are presented as described in the previous paragraph, however the CS+ is not paired with the US anymore. Furthermore, the background image is changed, representing a different context during extinction learning. On the following day, an extinction recall task is performed outside of the scanner, in order to assess the strength of the extinguished memory. The background image is the same as during fear extinction and the CS+ and CS- are presented without the administration of electrical stimulations (i.e., the US). A fear renewal task follows, to assess the strength of the fear memory in the context it was acquired in. The background image is therefore the same as the one used during the fear conditioning task. The CS+ and CS- are presented as described in the fear conditioning task, however without administration of electrical stimulations.

#### Explicit, exogenous stress-regulation – Emotional conflict resolution task

We will explore explicit, exogenous regulation by assessing behavioural responses and brain activity during a Stroop-like emotional conflict resolution task as described by Etkin et al. (2006). This task requires explicit attentional control and response inhibition over emotional processing that arises from conflicting external emotional information. The task consists of 148 presentations of happy or fearful facial expression photographs chosen from the set of Ekman and Friesen (1976). Faces are cropped and the words ‘‘FEARFUL’’ or ‘‘HAPPY’’ are written across the face in Dutch (i.e. “BANG”, “BLIJ”), such that word and expression are either congruent or incongruent. Stimuli are presented for 1s, with a varying interstimulus interval of 3– 5s, during which a central fixation cross is shown. Participants are instructed to respond as fast and accurately as possible, by selecting the response buttons corresponding to ‘‘fearful’’ or ‘‘happy’’ for the expression on the face. This task is designed to probe emotional conflicts by manipulating the congruency of the stimuli. Congruency refers to whether the two types of stimuli shown simultaneously in this experiment (i.e., facial expression and written word) match or not. For example, when a facial expression matches the word superimposed on it is considered a congruent trial, and when the facial expression does not match the word superimposed on it is considered an incongruent trial.

#### Implicit, endogenous stress-regulation - Resting under stress

We will assess resting-state functional connectivity within and between large-scale brain networks (ECN, SN, DMN) before and after a stress induction task, in order to investigate implicit, endogenous stress regulation. This task is chosen since it involves measuring spontaneous brain activity that is driven by an internal stressful state following a stress induction. Participants first undergo a resting state fMRI scan, for which they are instructed to keep their eyes open and fixated on a fixation cross on the screen. They are instructed to let their mind wander, without thinking of anything specific, or performing any repetitive mental activities. Shortly after this scan, a stress induction procedure takes place. The procedure is a modified version of the socially evaluated cold pressor test (SECPT), during which a researcher unknown to the participant will enter the MRI lab to perform the stress induction tasks. The researcher asks the participant to immerse their foot in a box of cold water (1.8-2.2 ° C) and try to keep it in for 3 minutes. The researcher then tells the participant that they are evaluating their facial expressions during the task. The researcher is instructed to abstain from giving positive feedback at all times and to try to keep a neutral facial expression. Once the Cold Pressor Test is over, the participants foot is covered with a towel and the researcher continues with the second part of the stress induction. In this part the participant has to count backwards, in steps of 17 or 13 from the number 1872, or 2013 respectively. The researcher asks them to be as fast and as accurate as possible. If they are too slow the researcher asks them to speed up. If they are incorrect the researcher asks them to start from the beginning. This task also lasts 3 minutes. After stress induction, a second resting-state fMRI scan is acquired, with the same instructions as before.

#### Explicit, endogenous stress-regulation – Self-regulation of brain networks under stress

A real-time fMRI neurofeedback paradigm, that has been recently developed and tested (54) is used to assess endogenous, explicit stress regulation after the previously described SECPT procedure. This task was selected as it requires active regulation of brain networks as a response to an internal stressful state. This task takes place after the SECPT and a resting state scan, well within the acute stress phase of the preceding stress induction. The stimulus used for neurofeedback presentation is a circle with a grey disc covering half of its area. Participants are asked to always fixate at the dot in the centre of the circle. During “Rest” the colour of the dot is black, and participants are instructed to rest, and think of nothing specific. During “Regulation” the dot is green, and arrows are pointing towards the outer circle or towards the centre of the circle, and participants are asked to perform the regulation task described below. The participants are instructed to control their brain activity, in order to change the size of the disc on the screen. They are told that they could do this by thinking of something specific, performing some mental task internally, or getting into a certain mood, emotion, feeling, or state of mind.

Participants are also instructed to try to avoid movement, including facial movements, limb movements and irregular breathing patterns. During “Feedback” the colour of the dot is orange, and the size of the grey disc will change reflecting the participant’s performance on the preceding regulation trial. The size of the disc directly reflects activity balance between the SN and ECN, therefore participants will have to learn for instance that increasing the size of the circle is possible by shifting the balance towards SN, while decreasing it is possible by shifting the balance towards ECN. Network balance is operationalised as the difference of the averaged activity of each of these two networks.

### Ecological Momentary Assessments (EMA)

The EMA consists of 6 days in which 6 surveys are filled throughout the day assessing mood, subjective stress levels, event appraisal, in combination with physiological monitoring for signs of stress (55,56). On all days of the week, participants are expected to fill in 6 short questionnaires on a smartphone app (SEMA3: http://www.sema3.com, Melbourne, Australia) that take maximally 3 minutes each to fill in. The first questionnaire of the day will additionally have questions regarding sleep quality, while the last will have items of self-reflection regarding the previous and the upcoming day. Additionally, during this week participants will wear an E4 wristband (Empatica sarl, Milano, Italy) measuring skin conductance, heart rate, skin temperature, and movement.

### Procedures

#### Recruitment

We recruit students via flyers on university campus sites, study advisors’ offices, student associations, and social media groups. Flyers will point students to our website which contains a more detailed description of our study and some necessary information for the procedures that we use. Interested individuals will find a link to a screening survey implemented on a platform specialised for anonymous recruitment of participants for scientific research (https:/www.soscisurvey.de). On this website, potential participants are asked to answer questions pertaining to the exclusion criteria described earlier, as well as the Perceived Stress Scale questionnaire.

Personal information and questionnaire scores are not accessible to the researchers. If all inclusion criteria and none of the exclusion criteria are met, potential participants are prompted to contact us. Thus, we are able to approach a large number of students and perform a pre-screening procedure that ensures their privacy. Once a potential participant contacts us, they receive detailed information about our study via email, and an appointment is scheduled. During this meeting, exclusion criteria are again checked, all measures, tasks, and procedures of the study are discussed, and an informed consent form is signed. Once participants join the study measurement sessions are planned, and participants receive regular email reminders to safeguard completion of the assessments and participant retention.

#### Baseline Measurements

After inclusion, the participant’s baseline measurements are acquired. First, two visits (on consecutive days) at the Donders Centre for Cognitive Neuroimaging (DCCN) are scheduled. On the first visit participants are asked to perform some of the neurocognitive assessments (Fear Conditioning task, Emotional Conflict Resolution task, and Resting State fMRI task). On the second visit participants are asked to perform a short task, part of the Fear Conditioning task. In addition, they undergo clinical assessments in the form of questionnaires on both days. Participants will then follow a week of ecological momentary assessments, during which they are asked to answer multiple short questionnaires during their daily life. After these baseline measurements, participants are randomly assigned to two groups as described earlier.

#### Intervention

Participants in the MBSR group receive the MBSR training and their attendance is logged, while participants in the wait-list group will not receive any planned intervention or measurements for the corresponding 8 weeks.

#### Post-intervention

As soon as the MBSR training has concluded, all participants are asked to visit the DCCN for their post-treatment measurements. These visits (visit 3 & 4) are scheduled on consecutive days and will involve the same neurocognitive and clinical assessments as the baseline measurements, with the addition of the Real-time fMRI Neurofeedback task. In addition, another ecological momentary assessment week is planned.

#### Follow-up

Two months later, during visit 5, clinical assessments are planned as a follow-up measurement. Participants in the wait-list group will then be able to receive the MBSR training, after which they are asked to visit the DCCN for a last clinical assessment.

### Statistical Analysis

#### Sample size calculation

The sample size, which is reported on http://trialregister.gov, was calculated on the basis of our planned Analysis of Covariance (ANCOVA) to estimate the difference between groups in perceived stress at post-treatment, while taking into account individual baseline measurements of perceived stress as a covariate. We use the method of Borm, Fransen, & Lemmens (2007), which adjusts the sample size calculated for an independent sample t-test by multiplying with the factor (1-r^2), where r is the correlation coefficient between baseline and post-treatment measurement. We aim to achieve a power of 80% with an *α=*0.05. The correlation between baseline and end-of-treatment perceived stress is estimated at *r=*0.7, based on data from Verweij et al. (2018). Assuming a moderate effect size of Cohen’s *d=*0.4, as reported by a large RCT on a general student sample (Galante et al., 2018), we would need 54 participants per group. Taking also into account an average attrition rate (10%) of studies on student populations reported in a meta-analysis by Khoury et al. (2015), we aim to recruit a total of 60 participants per group.

Notably, this estimate is conservative. We purposefully focused on one of the latest studies with a student population. The meta-analysis by Khoury et al. (2015) reports a higher combined effect size (*d=* 0.47) in 9 randomised controlled trials of student populations, on measures of stress, anxiety, depression, etc. This would result in our trial achieving a power of 91%. Additionally, this meta-analysis also suggests that the effect size of the treatment effect on different measures of perceived stress specifically is considerably higher than when combined with other outcomes mentioned above. The combined effect size estimate from nine studies on various healthy populations is *d=*0.74, which would lead to a power of 99% in our trial.

Sample size estimations based on the outcomes of our stress regulation tasks have proven to be exceptionally difficult due to the paucity of studies looking at the effects of MBSR on these specific tasks. However, there are indications that the influence of MBSR on some of these tasks is higher than the effect on clinical outcomes. For instance, large effect sizes are reported on extinction retention, the primary outcome of the fear conditioning and extinction task (*d=*0.93-1.15) (57,58). Studies assessing the effects of MBSR on resting state fMRI connectivity between regions within our networks of interest have reported a wide range of effect sizes (*d=*0.28-0.92) (59,60). Therefore, we expect to be able to detect effects of MBSR on these tasks with a well powered study based on our clinical outcomes.

#### Clinical symptoms

We expect that participants who follow the MBSR training will show a larger decrease in perceived stress, anxiety, and depressive symptoms compared to wait- list control participants. To statistically test the difference in perceived stress between the MBSR and the wait-list control groups post-treatment we will use an ANCOVA, including baseline PSS scores as covariates. Thus, the dependent variable will be post-treatment PSS score and group will be used as a between subject factor with two levels (MBSR, wait-list). The primary analysis will be performed with an intention-to-treat approach and sensitivity analyses will be conducted with different scenarios of imputed datasets to examine the influence of missing data on outcomes. Additional covariates will be added to account for individual differences (university, sex, age, and recruitment wave). Similar analyses will be performed on the following secondary clinical measures: depressive and anxiety symptoms, alcohol use, stress resilience, positive mental health and daily-life stress. Perceived stress at follow-up will be assessed by a linear mixed effect regression analysis with group (MBSR, wait-list), time (baseline, post-treatment), university, sex, age, and recruitment wave as fixed factors, as well as a group x time interaction. The intercept will be allowed to vary across participants to account for between-subject variability. Other clinical assessments, such as those assessing repetitive negative thinking, cognitive reactivity, allowing of emotions, mindfulness skills, self-compassion will be tested as potential mediating factors to determine whether changes in those scores after treatment can explain changes in the primary outcome at follow-up (i.e., PSS score). Mediation analyses will be conducted on the per-protocol sample (i.e., those participants who attended 50% or more of the training). Clinical assessments such as those assessing childhood trauma and personality traits at baseline will be tested as potential moderators of the clinical outcome.

#### MRI analysis

Neuroimaging data from all tasks will be analysed using various neuroimaging software packages, including FSL, SPM, (Turbo-) Brainvoyager, Nipype, as well as custom analysis scripts. The data will be first pre-processed, to remove artefacts, such as movement and physiological noise. General Linear Models will then be used per participant, per task, with regressors representing task activity, as well as nuisance parameters such as motion parameters and physiological noise parameters. Parameter estimates will then be introduced into second level a mixed-effect model with group (MBSR, wait-list), and time (baseline, post-treatment) as fixed factors, as well as a group x time interaction. Additional fixed factors will be added (university, sex, age, and recruitment wave). A random intercept for participants will be included to account for individual differences. In all tasks we will assess brain connectivity patterns of three large-scale brain networks (SN, ECN, DMN). Connectivity will be operationalised as the correlation coefficients between activity time-courses of nodes of these networks. Within- and between-network connectivity will be calculated to determine network cohesion and between-network communication, respectively. Potential connectivity patterns we observe (e.g., changes in SN or ECN within-network connectivity) will be tested as potential moderators of the clinical outcome post-treatment.

#### Implicit, exogenous stress-regulation – Fear Conditioning and Extinction task

Based on current fear learning and extinction literature we hypothesise that MBSR will enhance CS differentiation because of clinically promoted awareness to bodily signals and negative affect. We expect this heightened awareness during contextual extinction to lead to better extinction retention at day 2 without affecting fear renewal. Therefore, we expect less spontaneous recovery of the fearful memory when being exposed to the extinguished CS+ compared to the CS-. At the physiological level this means that smaller pupil diameter and skin conductance response differences between CS+ and CS- are expected in the MBSR group in the early stages of extinction retention compared to the wait-list control group.

Similarly, for our self-report measurements, we expect to observe lower perceived fear ratings in the MBSR group in the early stages of extinction retention, differentiating the CS+ and CS- compared to the wait-list control group. In our brain imaging measures, we expect that MBSR induces increased connectivity within SN during the extinction phase. We will statistically test differences in outcome physiological measures (skin conductance and pupillometry) between groups with mixed-effect regression analysis on each of the phases of the paradigm (conditioning, extinction, extinction retention, fear renewal). Fixed effects will include stimulus type (CS+, CS-), group (MBSR, wait-list), and time (baseline, post-treatment), as well as a group x stimulus type x time interaction. Additional fixed factors will be added (university, sex, age, and recruitment wave) and a random intercept for participants will be included to account for individual differences. Our main outcome in this task is extinction memory retention a day after extinction learning. Therefore, we will focus on the interaction effect of group x stimulus type x time during the extinction retention phase.

#### Explicit, exogenous stress-regulation – Emotional Stroop task

We expect that MBSR will affect participants’ responses when they are asked to identify and respond to emotionally conflicting information. In terms of behaviour, we expect to observe this as a reduction in response time in incongruent emotional situations. On our brain imaging measurements, we expect that MBSR will enhance ECN and SN within-network connectivity during emotional conflict processing. We will statistically test differences in behavioural outcome measures (reaction time and accuracy) between groups with linear mixed-effect regression analysis. Fixed factors will include group (MBSR, wait-list), time (baseline, post-treatment), congruency (congruent, incongruent), as well as a group x congruency x time interaction.

Additional fixed factors will be added (university, sex, age, and recruitment wave) and a random intercept for participants will be included to account for individual differences.

#### Implicit, endogenous stress-regulation - Resting state

Recent literature suggests that mindfulness enhances within-network connectivity of SN, decreases within-network cohesion of DMN, and decreases connectivity between SN and DMN (28). Furthermore, we expect that MBSR will additionally enhance within-network connectivity of both SN and ECN in the aftermath of exposure to an acute stressor, as well as enhancing connectivity between SN and ECN. To replicate mindfulness connectivity findings and test our hypothesis related to the effects of acute stress statistically, we will perform a network connectivity analysis as described earlier (MRI analysis) with an additional fixed factor determining pre-/post- stress induction data.

#### Explicit, endogenous stress-regulation – Neurofeedback task

We expect that the MBSR group will show a stronger self-regulation performance, compared to the wait-list group, due to an increased ability to dynamically change one’s large-scale brain network configuration. Self-regulation is defined as the ability to shift the SN-ECN network balance towards one or the other network at will. We expect that the MBSR group will be able to control the network balance by manipulating activity in their SN and ECN networks compared to the wait-list group who are expected to control the balance by manipulating activity mainly in their SN, as shown our recent proof-of-concept study (54). Self-regulation performance will be assessed using parameter estimates derived from a General Linear Model as described earlier. For this task the analysis will be done both online (i.e., while participants are in the MRI scanner), in order to provide feedback about their relative brain network configuration (54), and offline (i.e., after data acquisition).

Differences in self-regulation capability between groups will be tested offline using a linear mixed-effects regression analysis. Fixed factors will include group (MBSR, wait- list), university, sex, age, and recruitment wave and a random intercept for participants will be included to account for individual differences.

### Data Management

Handling of personal data is compliant with the EU General Data Protection Regulation. In all documents, subjects are identified by an identification code and access to personal data is granted only to members of the research team if necessary, and to the study monitor if requested. Saliva samples are destroyed after analysing their salivary cortisol content and that information is kept only in electronic form. All data are stored and archived through the data management infrastructure of the Donders Institute and will be kept for at least 15 years. It is estimated that there is no increased risk of harm when participating in this study.

Therefore, risk is negligible, and a data monitoring committee is not needed for this study.

## Discussion

### Summary

This study investigates the effect of MBSR on clinical measures in a student population with high self-reported levels of perceived stress in a longitudinal, randomised wait-list controlled trial. The current study aims to establish a better understanding of *how* MBSR changes one’s stress regulation. Outcomes on different levels of measurement, such as self-report, behavioural, physiological, and brain imaging will be assessed at different timepoints providing novel insights into the working mechanisms of MBSR. Moreover, from this mechanistic point of view we will explore whether we can uncover moderators of treatment effect, i.e., for whom this intervention might be more beneficial in terms of reduction of stress-related symptoms.

### Strengths

The use of a formal mindfulness intervention, such as MBSR, is a strength of this study. This makes the current study reproducible and comparable to other studies using the same formal intervention. Moreover, the tasks included cover a broad range of stress-regulation processes: Implicit/explicit and endogenous/exogenous stress-regulation is assessed using different measurement techniques, such as self-report, behavioural testing, physiological outcomes, and brain imaging, providing a very rich dataset. This study will be one of the largest MBSR randomised controlled trials to include neuroimaging measures. Our findings can positively contribute to the increasing literature of studies exploring neural mechanisms of mindfulness-based interventions. Finally, this is a longitudinal study, following participants’ stress-related clinical outcomes over a period of 7-10 months. This is key to determine the long-term effects of the MBSR training in a population susceptible to stress-related symptomatology.

### Limitations

The specific population chosen for this study (i.e., university students preselected on high perceived stress levels) limits generalisability to the general public, considering, e.g., the age and education range. In addition, all included tasks are designed specifically to evaluate acute stress reactivity on a relatively narrow time window.

This means that further experiments are necessary to investigate the impact of MBSR on stress recovery at a longer timescale. Furthermore, participant group allocation takes place after their baseline measurements, therefore these are blinded with respect to treatment allocation to the participants as well as the researchers. For pragmatic reasons, however, researchers performing the post-treatment measurements are also performing group allocation and participant communication and therefore post-treatment measurements are not blinded.

### Update

We are currently in the process of recruiting and testing participants since April 2021. It is expected that reaching the total sample size which was reported on trialregister.gov will be a challenge. One of the main factors contributing to this is the impact of the COVID-19 pandemic lockdowns, since during the first year of recruitment students were largely studying remotely. In order to address this issue, the study’s recruitment period has been extended by a year, and recruitment efforts have been reassessed. However, there is reason to believe that our sample size estimate is very conservative. The meta-analysis by Khoury et al. (2015) suggests that the combined effect size from studies investigating MBSR on student populations is higher than our original estimate. They report even higher combined effect size on stress measures in studies with various healthy populations, including students. This suggests that our trial is better powered than initially thought. In addition, effect sizes reported in studies investigating the effects of MBSR on stress regulation tasks, similar to the ones we deploy, suggest that the current trial will be well powered to detect these effects (57–60). This makes us confident that we will be able to detect differences in perceived stress and our stress regulation tasks despite potentially reaching a smaller sample size.

## Conclusion

In conclusion, this study aims to provide clinicians and scientists alike with valuable insights into how effective MBSR is in reducing stress-related symptomatology in a susceptible student population, how it affects stress regulation in this population and for whom it might be more beneficial. While limitations exist, the use of a formal mindfulness intervention during a longitudinal study in combination with the use of a wide range of stress-regulation tasks make this study a valuable contribution to field of stress and mindfulness research. Moreover, by identifying characteristics of individuals who are most likely to benefit from MBSR, clinicians can effectively target those individuals, as well as tailor the treatment to fit one’s individual needs. A better understanding of how and for whom MBSR works best could lead to more future research and aid the development and implementation of MBSR programs.

## Data Availability

Scripts used for stimulus presentation, analysis scripts, and non-sensitive pseudonymised data such as questionnaire data, behavioural data, pre-processed physiology data, and group-level MR data will be openly available in the Open Science Framework at https://osf.io. Pseudonymised MR images will be saved on the Donders Repository at https://data.donders.ru.nl and will be made available upon request to the corresponding author. Raw MR images will not be available due to privacy restrictions. Trial results will be disseminated in relevant peer-reviewed journals and scientific conferences.

## Abbreviations

MBSR: Mindfulness Based Stress reduction,
MBI: Mindfulness Based Interventions,
ACC: Anterior Cingulate Cortex,
PFC: Pre-frontal Cortex
SN: Salience Network,
DMN: Default Mode Network,
ECN: Executive Control Network,
PSS: Perceived Stress Scale,
IDS-SR: Inventory of Depressive Symptomatology Self-Report
STAI: State and Trait Anxiety Inventory,
AUDIT: Alcohol Use Disorders Identification Test,
MACE-X: Maltreatment and Abuse Chronology of Exposure Scale
NEO-FFI: NEO Five Factor Inventory,
PTQ: Perseverative Thinking Questionnaire,
LEIDS-R: Leiden Index of Depression Sensitivity – Revised
AAQ: Acceptance and Action Questionnaire,
FFMQ-SF: Five-Facet Mindfulness Questionnaire
SCS-S: Self-Compassion Scale,
CD-RISK: Connor-Davidson Resilience Scale
MHC-SF: Mental Health Continuum,
fMRI: Functional Magnetic Resonance Imaging,
SECPT: Socially Evaluated Cold Pressor Test
EMA: Ecological Momentary Assessments,
DCCN: Donders Centre for Cognitive Neuroimaging
ANCOVA: Analysis of Covariance,
RCT: Randomised Controlled Trial

## Declarations

## Acknowledgements

Not applicable.

## Author contributions

EH, AS, FK, and DG designed the study and wrote the grant application. NK, DG, and FK drafted the first versions of the manuscript, which was modified and supplemented by EH, AS. EH and AS are the principal investigators and AS is responsible for supervision of the mindfulness trainers. NK is involved in recruiting participants, obtaining participants’ informed consent, and is responsible for the logistics and data collection. All authors have read and approved the final paper.

## Availability of data and materials

Scripts used for stimulus presentation, analysis scripts, and non-sensitive pseudonymised data such as questionnaire data, behavioural data, pre-processed physiology data, and group-level MR data will be openly available in the Open Science Framework at https://osf.io. Pseudonymised MR images will be saved on the

Donders Repository at https://data.donders.ru.nl and will be made available upon request to the corresponding author. Raw MR images will not be available due to privacy restrictions. Trial results will be disseminated in relevant peer-reviewed journals and scientific conferences.

## Ethics approval and consent to participate

The MindRest trial has been approved by the local medical ethics committee, METC Oost-Nederland (number: NL74345.091.20). The study is conducted according to the principles of the Declaration of Helsinki (6th edition, 2008) and in accordance with the Medical Research Involving Human Subjects Act (WMO). Participation is voluntary and a written informed consent is obtained from all the participants. We believe that the risks of participation are negligible. However, we will monitor for potential adverse events. Any significant modifications to the protocol will be submitted as an amendment to the medical ethics committee.

## Consent to publish

Not applicable.

